# Cognitive impairments in sporadic cerebral small vessel disease (SVD): a systematic review and meta-analysis of cohorts with stroke, dementia and non-clinical presentations of SVD

**DOI:** 10.1101/2020.02.10.20020628

**Authors:** Olivia KL Hamilton, Ellen V Backhouse, Esther Janssen, Angela CC Jochems, Caragh Maher, Anna J Stevenson, Tuula E Ritakari, Lihua Xia, Ian J Deary, Joanna M Wardlaw

**Affiliations:** Centre for Clinical Brain Sciences, University of Edinburgh, Chancellor’s Building, 49 Little France Crescent, Edinburgh, UK, EH16 4SB; Lothian Birth Cohorts, University of Edinburgh, 7 George Square, Edinburgh, UK, EH8 9JZ; Centre for Genomic and Experimental Medicine, MRC Institute of Genetics and Molecular Medicine, University of Edinburgh, Western General Hospital Campus, Crewe Road, Edinburgh, UK, EH4 2XU; Centre for Discovery Brain Sciences, University of Edinburgh, Hugh Robson Building, 15 George Square, Edinburgh, UK, EH8 9XD; Edinburgh Dementia Research Centre in the UK Dementia Research Institute, Chancellor’s Building, 49 Little France Crescent, Edinburgh, UK, EH16 4SB; Department of Psychology, University of Edinburgh, 7 George Square, Edinburgh, UK, EH8 9JZ

## Abstract

**Background:** Cognitive impairment is a key clinical feature of cerebral small vessel disease (SVD), but the full range of SVD-related cognitive impairments is unclear, and little is known about how they might vary across clinical and non-clinical manifestations of SVD.

**Methods:** In systematic searches of OVID MEDLINE, Embase, and PsychINFO from 1^st^ January 1985 to 6^th^ October 2019, we identified studies reporting cognitive test results for study participants with SVD and control participants without SVD. Using standardised group-level cognitive test data, we performed random effects meta-analyses in seven cognitive domains to test whether cognitive test scores differed between SVD and control groups. We conducted meta-regression analyses to test whether differences in age, education, or vascular risk factors between SVD and control groups, or whether different clinical manifestations of SVD (e.g. stroke, cognitive impairment, or non-clinical presentations) accounted for cognitive effect sizes.

**Findings:** Of 8562 studies identified, we included 69 studies from six continents, published in four languages. These studies included 3229 participants with SVD and 3679 controls. Meta-analyses demonstrated that on average, control groups outperformed SVD cohorts on cognitive tests in all cognitive domains examined: executive function (estimate: -0.928; 95%CI: -1.08, -0.78); processing speed (-0.885; -1.17, -0.60); delayed memory (-0.898; -1.10, -0.69); language (-0.808; -1.01, -0.60); visuospatial ability (-0.720; -0.96, -0.48); reasoning (-0.634; -0.93, -0.34); and attention (-0.622; -0.94, -0.31; all p≤0.001). Meta-regression analyses suggested that differences in years of education between SVD and control groups may account for a proportion of the differences in performance on tests of executive function, visuospatial ability and language, and that cohorts with cognitive impairments performed more poorly on tests of executive function, delayed memory and visuospatial ability than cohorts with stroke or non-clinical presentations of SVD.

**Interpretation:** Participants with SVD demonstrated poorer cognitive performance relative to control groups in all cognitive domains we examined. This effect was present for all presentations of SVD, reinforcing the need to test a range of cognitive domains in both clinical and research settings. Lower levels of education in SVD versus control participants may contribute to these effects, highlighting the need to account for educational level in the assessment of SVD-related cognitive impairment.

**Funding:** None.

## Introduction

Cerebral small vessel disease (SVD) refers to a collection of neuroimaging and neuropathological abnormalities found in the brain’s white and deep grey matter. White matter hyperintensities and lacunes of presumed vascular origin, cerebral microbleeds and enlarged perivascular spaces likely reflect multiple pathological changes affecting the brain’s small vessels, such as endothelial dysfunction, impaired cerebral blood flow, and reduced vessel pulsatility. The relationships between these mechanisms are complex and not yet fully understood^1,2^. Whereas the radiological markers of SVD have previously been considered clinically ‘silent’, their presence has been associated with cognitive impairment, depression, impaired gait and balance, and urinary incontinence^3,4^.

SVD has varied clinical manifestations - it causes approximately 25% of acute ischaemic stroke, increases the risk of recurrent ischaemic stroke, and associates with poorer functional outcomes post-stroke^5,6^. SVD is also a major cause of vascular dementia (VaD), and increases the risk of incident dementia^6^. In most cases, SVD manifests sub-clinically, with few overt symptoms. For example, cohort studies of healthy older individuals often include subpopulations who show features of SVD on neuroimaging, but have no history of stroke, dementia, and no subjective cognitive concerns. The term vascular cognitive impairment (VCI) encompasses these various presentations of SVD, referring to any severity of cognitive impairment (from subclinical deficits to dementia) with underlying vascular contributions^7,8^.

The cognitive manifestation of VCI is often characterised as deficits in executive function and speed of information processing, with little attention paid to other domains of cognitive ability^9,10^. Studies supporting this suggestion are often small and focus on a narrowly-defined subtype of SVD, or on those with a high disease burden, who may not represent the full spectrum of sporadic SVDs. Despite increasing recognition of cerebrovascular contributions to non-vascular and mixed dementias^11^, individuals with cerebrovascular presentations of SVD are rarely considered in the same study as those with predominantly cognitive presentations, so little is known about how cognitive impairments may differ between SVD subtypes. In part, this is due to differing routes into clinical care and so, into clinical research. In addition to this, variation in naming conventions for SVD lesions^12^ impairs between-study comparisons.

To examine the nature of SVD-related cognitive impairment across multiple cognitive domains and to account for its varied clinical and non-clinical presentations, we conducted a systematic review and meta-analysis of domain-specific cognitive abilities in individuals with clinical or radiological signs of SVD. We aimed to clarify the nature of SVD-related cognitive impairment, to assess contributions of underlying factors such as age, level of education or burden of vascular risk, and to assess whether SVD-related cognitive impairments vary according to clinical, or non-clinical presentations of the disease.

## Methods

We performed this systematic review and meta-analysis in accordance with PRISMA guidelines. The review protocol is registered on the PROSPERO database (registration number: CRD42017080215).

### Search strategy and study selection criteria

We developed and tested a detailed search strategy (Appendix A) to identify studies reporting the results of cognitive testing in a cohort with SVD (performed contemporaneous with identification of SVD), and a control cohort with no history of neurological or psychiatric conditions. We searched OVID MEDLINE, Embase and PsychINFO, for human studies published in any language from 1^st^ January 1985, when MRI became more widely available in clinical practice, to 6^th^ October 2019. To identify additional studies, we checked the reference lists of relevant review articles and hand-searched the previous 7 years of *Stroke* and the *Journal of Cerebral Blood Flow and Metabolism*. Study inclusion and exclusion criteria are presented in Appendix B.

### Data Analysis

Two authors (OH and EB) independently extracted the following information: country in which the study was carried out; recruitment setting; study inclusion/exclusion criteria; clinical diagnosis or other descriptive characterisation of the SVD cohort(s) (including duration of symptoms/diagnosis and diagnostic criteria); sample size of the SVD and control groups; group-level demographic data for the SVD and control groups (age, sex, education); group-level data on vascular risk factors (% cohorts with hypertension, diabetes, hypercholesterolemia, and smoking status), group-level data on WMH burden, and group-level cognitive test scores for SVD and control groups. The vast majority of cognitive data were presented as mean and standard deviation. To avoid introducing additional heterogeneity into the meta-analysis dataset, we did not convert cognitive data presented as median and range to mean and standard deviation - instead these data are summarised in Appendix C. Where individual participant data were presented, we calculated the mean and standard deviation of the variables we extracted.

Two authors (OH and AJ) independently grouped cognitive data into seven domains: information processing speed, executive function, delayed memory, attention, reasoning, visuospatial ability and language, and resolved disagreements by consensus. It is important to note that subdomains of cognitive ability are not discrete, and that individual cognitive tests often engage abilities across multiple domains – for transparency, test groupings provided in Appendix D. Studies reported a wide range of memory tests, including tests of long-term, short-term and working memory. To reduce heterogeneity in the dataset we included only tasks featuring a delayed recall/recognition component, as these were the most frequently reported memory tasks. We excluded data for which we could not identify the specific test score (e.g. where authors reported results for a Trail Making task, but did not specify whether the score was for Trail Making A, Trail Making B, or Trail Making A-B). We also excluded data for which we could not discern whether a higher or lower score indicated better performance. Where studies reported multiple scores for one cognitive test (e.g. for the Wisconsin Card Sorting Test: number of perseverative errors, total number of errors, number of categories etc.), we included the score most commonly reported in the meta-analysis dataset.

### Effect sizes

We calculated a standardised mean difference to represent the difference between performance of the SVD and control cohorts on each cognitive test. We multiplied the SMD by -1 for tests on which a lower score indicated better performance.

### Meta-analysis models

We ran seven separate random effects meta-analyses to assess the difference in performance between SVD and control groups on cognitive tests in each cognitive domain. We conducted all meta-analyses using the robumeta package^13^ in R version 3.6.1^14^. robumeta permits the meta-analysis of multiple effect sizes from one study by employing robust variance estimation (RVE) to account for their statistical dependency. This approach maximises the amount of data derived from a single study, increasing the statistical power of each meta-analysis. Dependency in our dataset arose from the inclusion of multiple effect sizes from the same study sample, and the inclusion of studies which used the same control group for comparison with multiple SVD groups. Covariance matrices for multiple outcomes arising from a single study are rarely published, therefore, robumeta imputes a user-specified value for the within-study effect size correlation. We were conservative in our choice of within-study effect size correlation - we specified rho as 0.8 and carried out sensitivity analyses in robumeta, which impute rho values at increments of 0.1 to test whether this alters the model results. For all analyses, we weighted effect sizes according to a correlated effects dependence structure within the robumeta package and used small sample size corrections. Small sample corrections, which correct both the residuals and the degrees of freedom (df) used in the RVE, increase the accuracy of models including less than 40 studies^13^. After correction, if the Satterthwaite df for the model are less than four, the p value is considered unreliable due to the probability of type I error being greater than 0.05. In the proceeding analyses, results of models with Satterthwaite df<4 were considered unreliable. We report I^2^ and τ^2^ as measures of heterogeneity.

### Meta-regression models

We carried out two secondary analyses to examine the following study-level and cohort-level variables:

#### 1) SVD presentation

To test whether the pooled study effect size differed according to SVD presentation, we grouped each SVD cohort into one of three categories below. We then entered SVD presentation as an ordinal predictor in the meta-regression model for each cognitive domain, with the cognitive impairment category acting as the reference group.

##### a) Predominantly cerebrovascular presentations of SVD

Cohorts in this group most commonly presented to clinical stroke or neurology services with symptoms of lacunar syndrome, with or without radiological evidence of corresponding vascular lesions. Other cohorts in this category had clinical diagnoses of SVD or subcortical ischaemic vascular disease.

##### b) Cognitive presentations of SVD

Cohorts in this group were identified on the basis of impaired cognitive ability ranging from mild cognitive impairment (MCI) to VaD. Typically, cohorts presented with cognitive impairment (according to clinical diagnosis, or objective cognitive testing) and either radiological evidence supporting a vascular aetiology, or multiple risk factors for cerebrovascular disease.

##### c) Non-clinical presentations of SVD

This group included cohorts with radiological evidence of SVD (WMH or lacunes of presumed vascular origin), but no clinical diagnosis. Typically, cohorts were community-dwelling older individuals recruited within a defined geographical region, or via community advertising. Several cohorts in this category presented to clinical services with non-specific symptoms such as dizziness or headache, but received no diagnosis upon examination.

#### 2) Differences in age, education, hypertension and diabetes between the SVD and control cohorts

All extracted cognitive data were raw scores, unadjusted for demographic or vascular risk factors. Therefore, to test whether differences in age, education, hypertension and diabetes between SVD and control cohorts accounted for study effect sizes, we calculated the difference in age, years of education, % sample with hypertension, and % sample with diabetes (e.g. difference in age = mean age of control cohort – mean age of SVD cohort), and entered these variables as predictors in separate univariate meta-regression models for each cognitive domain.

### Quality assessment

Quality assessment criteria (see Appendix E) were devised according to STROBE guidelines. Two authors (OH and EJ) independently assessed the quality of included publications and resolved disagreements by consensus. To assess whether the inclusion of lower quality studies affected the results of the meta-analyses, we re-ran meta-analysis models excluding studies with quality scores lower than the median quality score of the meta-analysis sample.

## Results

Our literature search identified 8562 studies (see figure 1). After screening titles and abstracts, the full texts of 1359 publications were assessed against inclusion and exclusion criteria. We extracted data from 69 studies^15–83^, which reported data for 89 cohorts with SVD (n=3229), and 71 control cohorts (n=3679). Characteristics of included studies are presented in table 1. Studies were conducted in 18 countries across six continents (Africa=1; Asia=31; Australia=1; Europe=18; North America=16; South America=2), and published in four languages.

**Figure 1:**
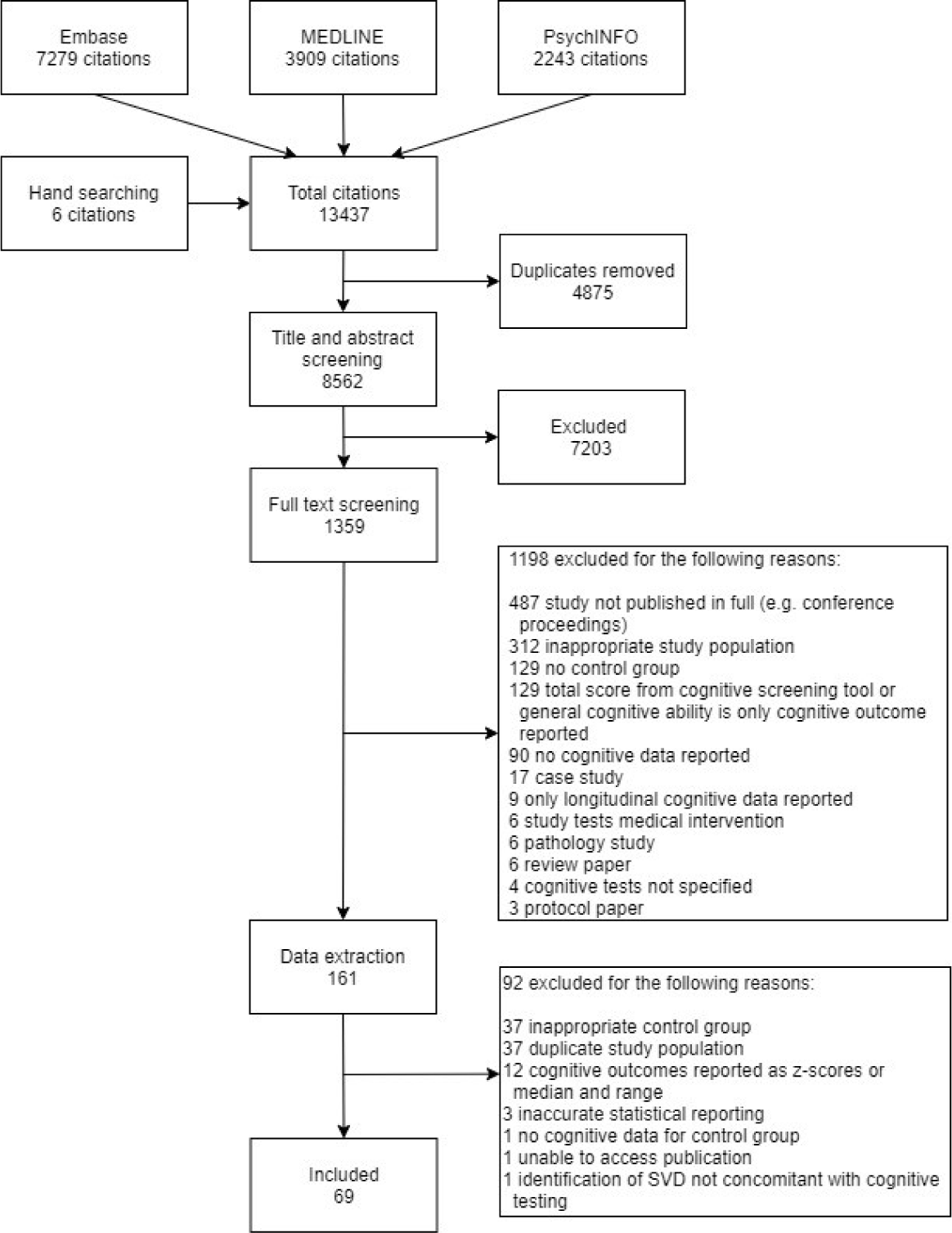
Study selection flow diagram

**Table 1:**
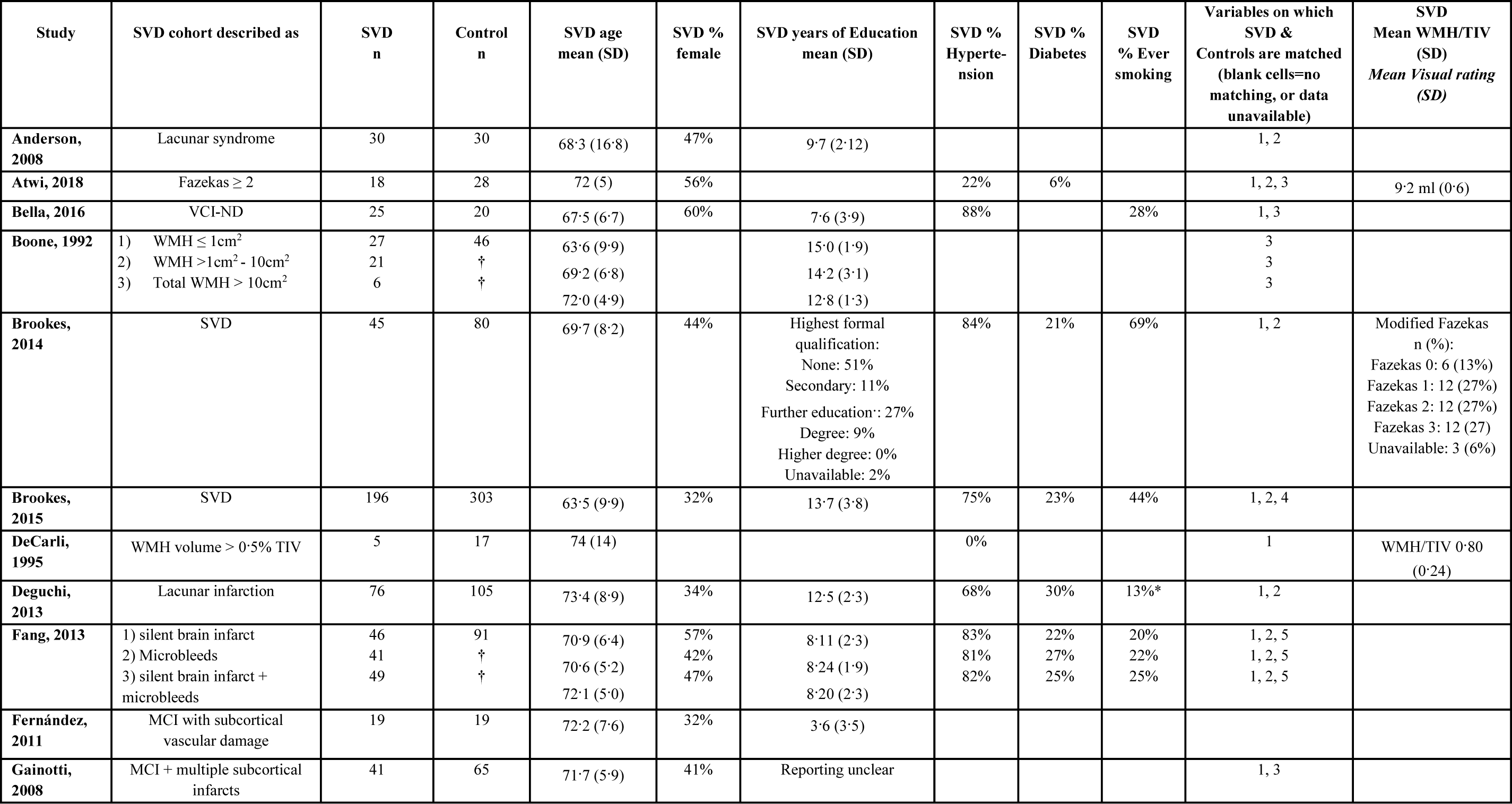
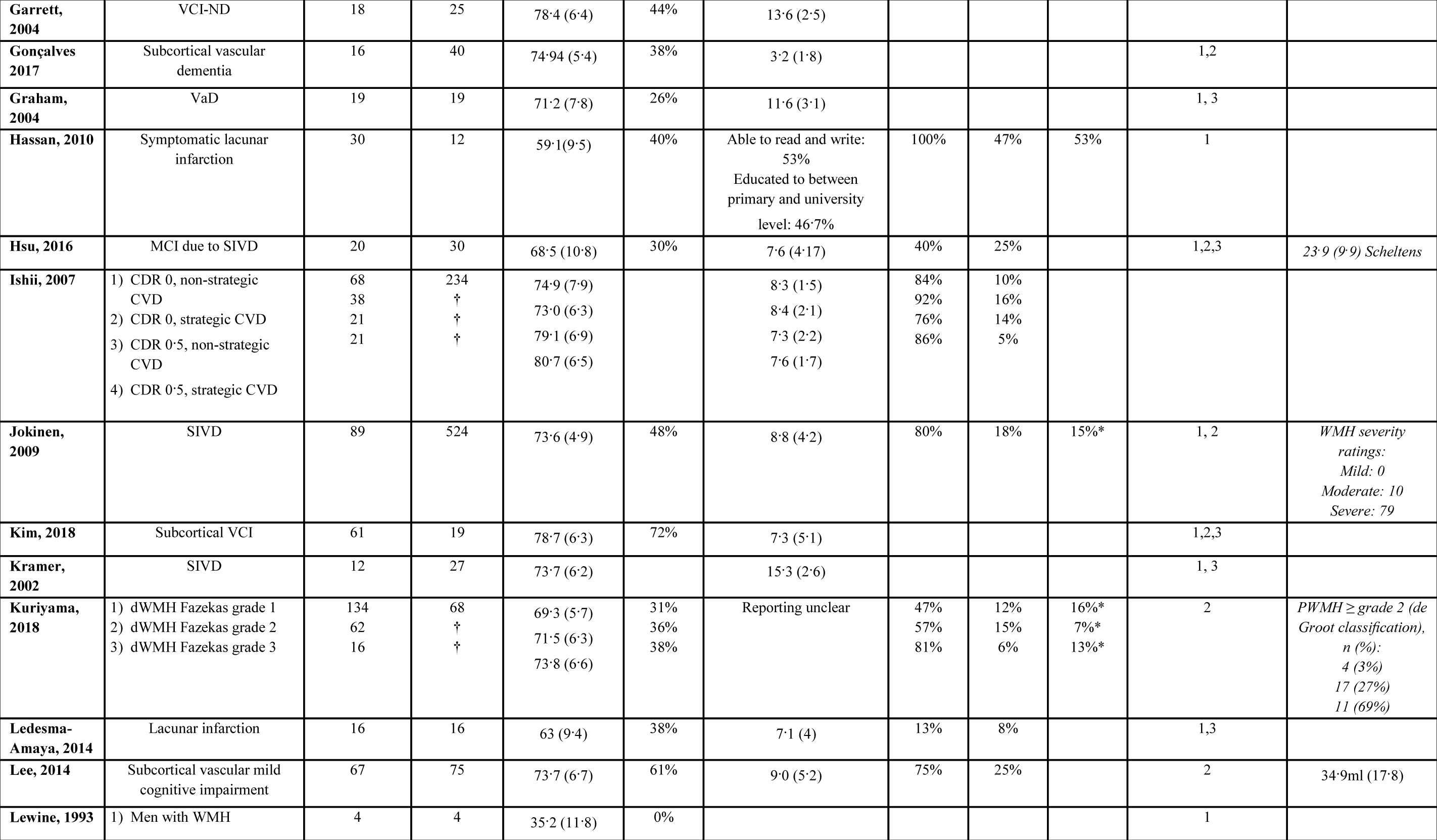
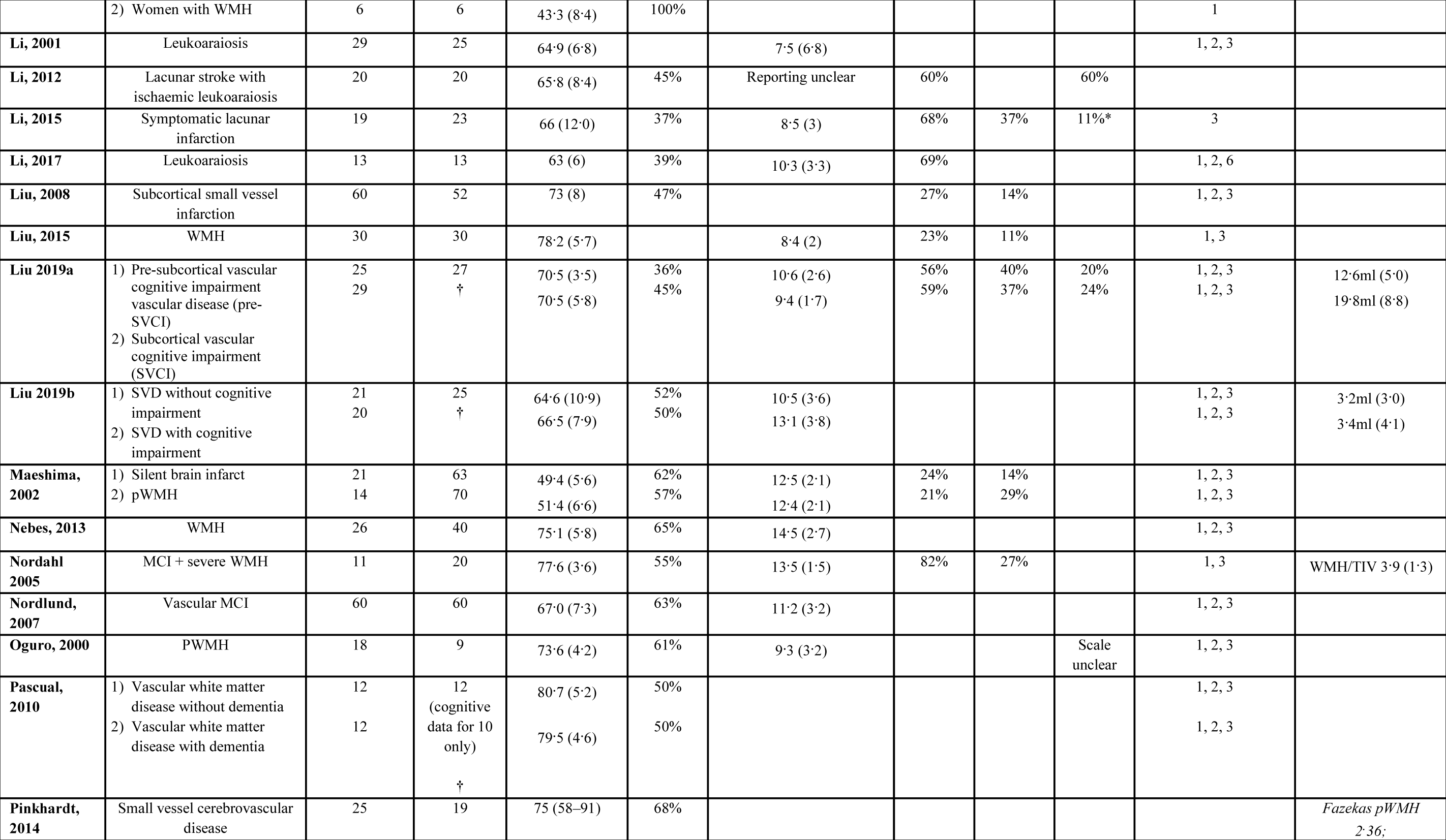
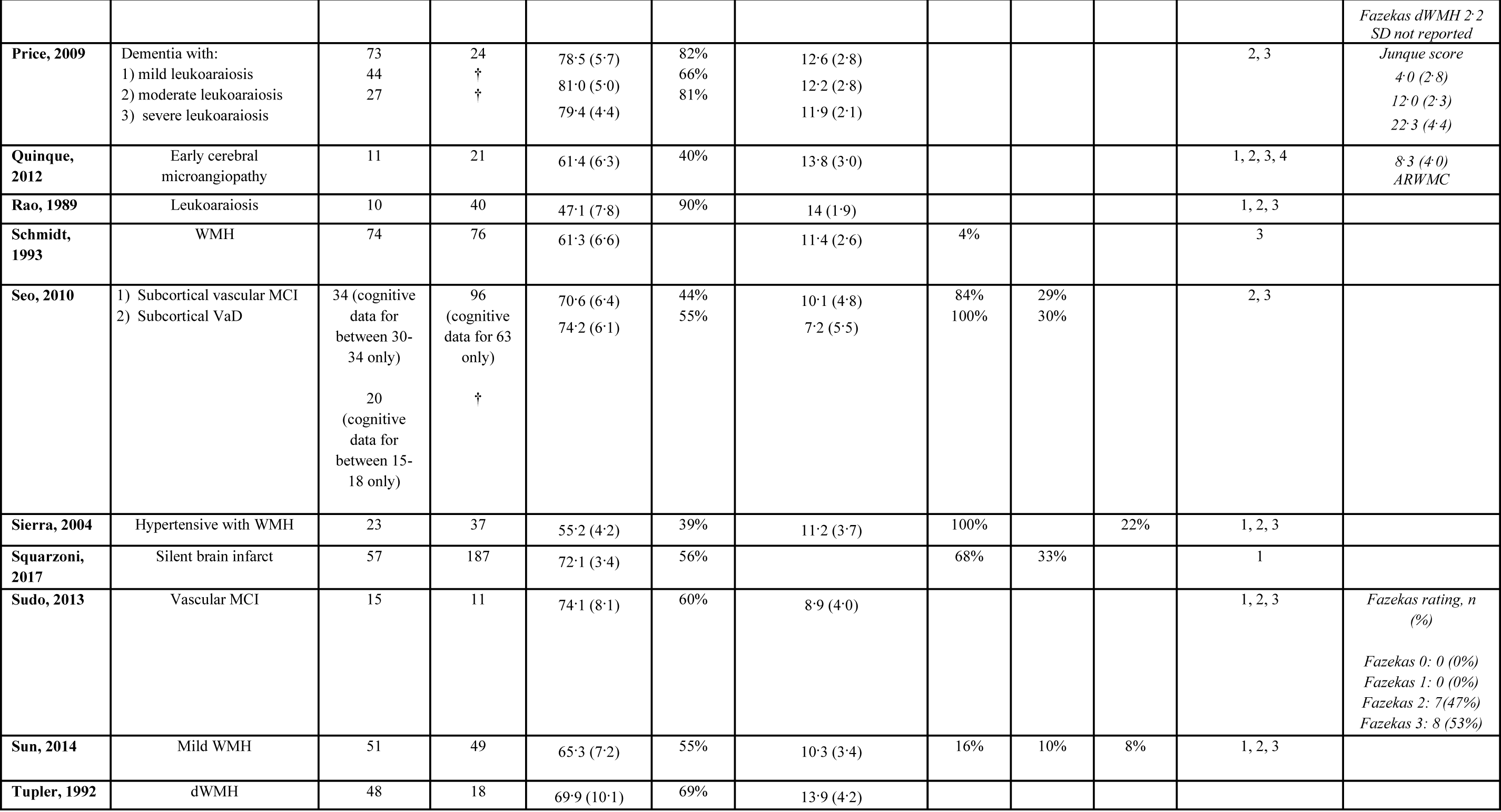
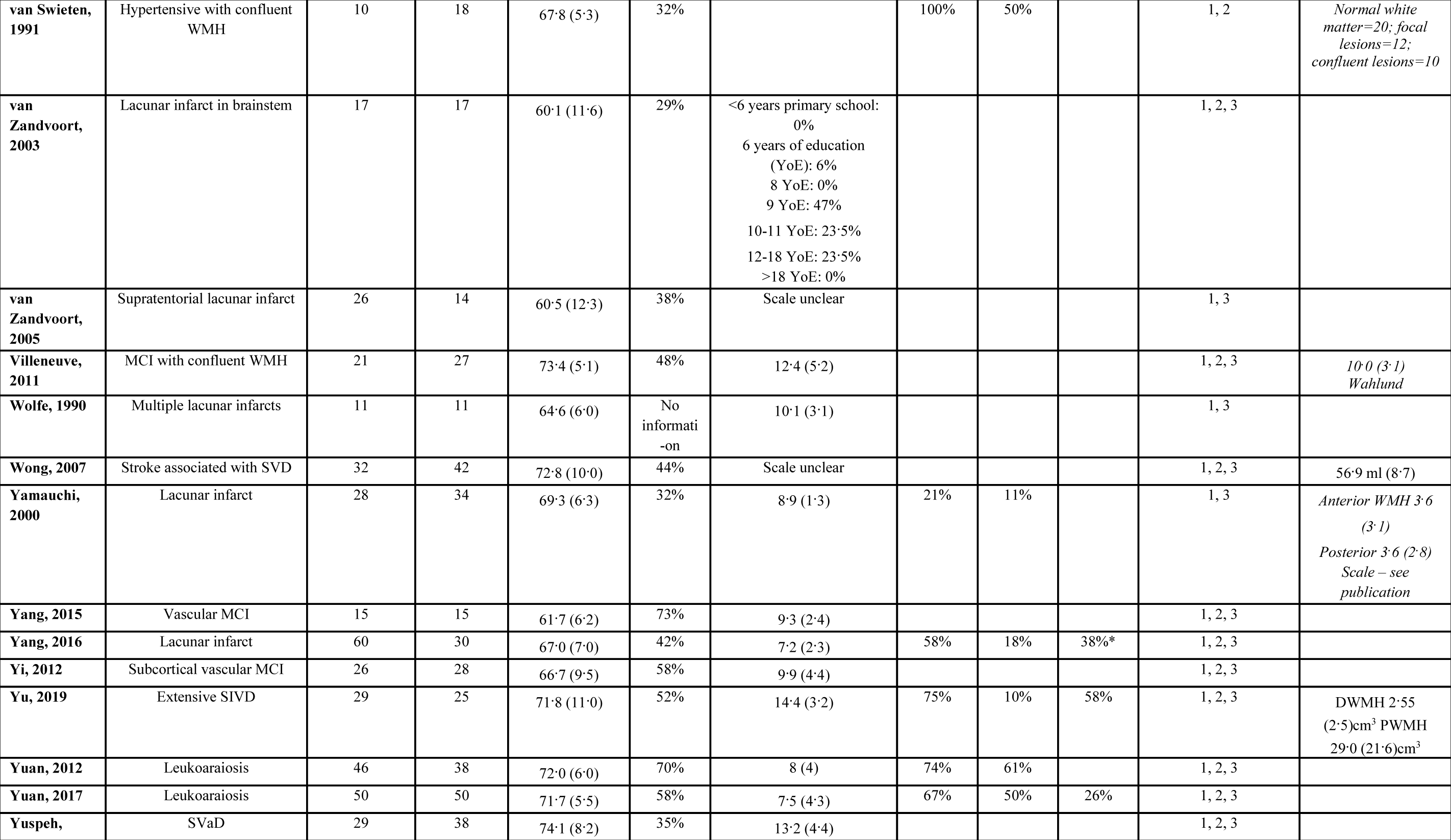
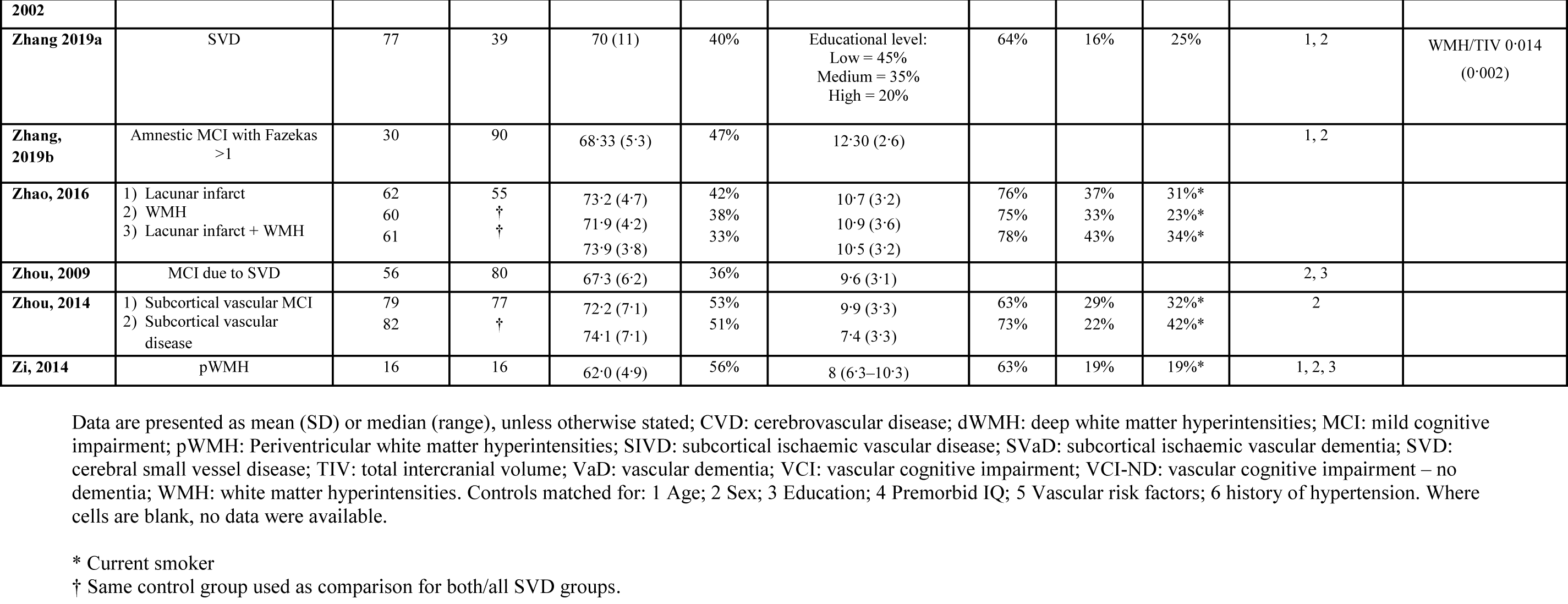
Characteristics of all included studies

Demographic data for the 89 SVD and 71 control cohorts are presented in table 2. We did not test for differences in age, sex, level of education, or vascular risk factors between the SVD and control groups as some studies only reported these data for the SVD group, therefore, comparisons would not include all participants contributing cognitive data to the meta-analyses. Almost all studies reported participants’ mean age and sex, but the reporting of educational level, vascular risk factors, and WMH burden was less complete (see Table S1).

**Table 2:** Demographics of SVD and control cohorts from all included studies

|  | SVD cohorts |  | Control cohorts |  |
| --- | --- | --- | --- | --- |
|  | cohorts (n=89) | mean (SD or 95% CI) | cohorts (n=71) | mean (SD or 95% CI) |
| mean age* | 88 | 69.3 (67.8, 70.9) | 70 | 66.4 (64.6, 68.2) |
| % female | 76 | 49.0 (15.9) | 63 | 50.9 (15.0) |
| mean years education* | 67 | 10.3 (9.7, 10.9) | 53 | 10.8 (10.1, 11.6) |
| % hypertension | 48 | 66.7 (23.0) | 34 | 37.8 (20.7) |
| % diabetes | 45 | 25.5 (13.7) | 31 | 17.1 (13.5) |
| % hypercholesterolemia | 5 | 55.1 (20.0) | 4 | 35.1 (12.3) |
| % history of smoking | 28 | 28.3 (16.1) | 16 | 25.6 (16.9) |

\* Mean age and mean years of education were calculated using random effects meta-analysis in the meta package<sup>1</sup> in R version 3.6.1. Only studies that presented group level data for age and years of education as mean and standard deviation were included in these meta-analyses.
1. Schwarzer G. “meta: An R package for meta-analysis.” R News 2007, 7(3), 40–45.

Tests of executive function were the most commonly reported cognitive outcomes (58 studies reported 188 cognitive outcomes), followed by tests of delayed memory (41 studies, 98 outcomes), processing speed (37 studies, 88 outcomes), visuospatial ability (27 studies, 50 outcomes), language (24 studies, 42 outcomes), reasoning (16 studies, 25 outcomes), and attention (12 studies, 19 outcomes; see table S2).

We performed meta-analyses to estimate the differences in performance on cognitive tests, between cohorts with SVD and control cohorts, in seven cognitive domains. A negative effect size estimate indicates that on average SVD cohorts performed more poorly on cognitive tasks than control cohorts, an effect size of zero indicates no difference between the two groups, and a positive effect size indicates that on average the control cohorts performed more poorly on cognitive tasks than SVD cohorts. The pooled estimated effect size for each meta-analysis model demonstrated that the control cohorts outperformed the SVD cohorts on cognitive tasks in all seven domains (see table 3, and supplementary figures S1 – S7). Sensitivity analyses suggested that pooled estimated effect sizes were robust to different within-study effect size correlations – varying the values of rho between 0 and 1 altered the SE of the pooled effect size and the τ^2^ estimate at the second or third decimal place. I^2^ values from the meta-analysis models were large, indicating a high proportion of inconsistency between effect sizes within each domain.

**Table 3:** Results of meta-analysis models for each cognitive domain

|  |  |  |  |  |  |  | **Heterogeneity** |  |
| --- | --- | --- | --- | --- | --- | --- | --- | --- |
| | **Studies** | **Outcomes** | **Estimate (SE)** | **95% CI** | **Degrees of freedom** | **Uncorrected p value** | $\tau^2$ | $I^2$ |
| Processing Speed | 37 | 88 | -0.885 (0.14) | -1.17, -0.60 | 35.8 | $2.3 \times 10^{-7}$ | 0.6 | 91.4 |
| Executive function | 58 | 188 | -0.928 (0.08) | -1.08, -0.78 | 56.2 | $< 2 \times 10^{-16}$ | 0.4 | 87.9 |
| Delayed memory | 41 | 98 | -0.898 (0.10) | -1.10, -0.69 | 39.6 | $7.2 \times 10^{-11}$ | 0.5 | 88.0 |
| Attention | 12 | 19 | -0.622 (0.14) | -0.94, -0.31 | 10.6 | 0.001 | 0.2 | 80.8 |
| Reasoning | 16 | 25 | -0.634 (0.14) | -0.93, -0.34 | 14.6 | $4.2 \times 10^{-4}$ | 0.2 | 76.5 |
| Visuospatial ability | 27 | 50 | -0.720 (0.11) | -0.96, -0.48 | 25.3 | $1.3 \times 10^{-6}$ | 0.3 | 77.6 |
| Language | 24 | 42 | -0.808 (0.10) | -1.01, -0.60 | 22.7 | $3.2 \times 10^{-8}$ | 0.3 | 81.2 |

The meta-analysis dataset included 26 cohorts with predominantly cerebrovascular presentations of SVD, 31 cohorts with cognitive presentations of SVD, and 32 cohorts with non-clinical presentations of SVD. Diagnostic and radiological criteria varied between studies – where available this information was extracted and is presented in Appendix F. Demographics of the three SVD presentation categories are presented in table S3. There were no differences in years of education, or prevalence of hypertension or diabetes between the three SVD presentation categories, but cohorts with a cognitive presentation of SVD were significantly older than those with non-clinical presentations of SVD (p=0.002). The results of meta-regression models assessing differences in cognitive performance between cohorts with cerebrovascular, cognitive and non-clinical presentations of SVD indicated that cohorts with a cognitive presentation of SVD performed worse on tests of delayed memory than cohorts with cerebrovascular and non-clinical presentations (see table S4). The effect size for delayed memory was 0.83 standard deviations greater for the cerebrovascular cohorts (95% CI: 0.44, 1.21; p<0.001) and 0.85 standard deviations greater for non-clinical cohorts (95% CI: 0.40, 1.29; p=0.001), than cohorts with a cognitive presentation of SVD. Cohorts with a cognitive presentation of SVD also performed more poorly on tests of executive function and visuospatial ability than the non-clinical cohorts. The effect size was 0.46 standard deviations greater in the domain of executive function (95% CI: 0.07, 0.86; p=0.023), and 0.68 standard deviations greater in the domain of visuospatial ability (95% CI: 0.30, 1.01; p=0.002) for the non-clinical cohorts than the cohorts with a cognitive presentation of SVD. Including SVD presentation as a predictor in meta-regression models had little effect on study heterogeneity. I^2^ values increased by 0.4% and 2.2% in the domains of processing speed and reasoning, but decreased by 5.5%, 5.9% and 11.2% in the domains of attention, memory and language, when compared to univariate meta-analysis models, and to a lesser extent in the domains of executive function and visuospatial ability. Overall, however, I^2^ values remained large.

We carried out further meta-regression analyses to test associations between differences in age, years of education, and prevalence of hypertension and diabetes between SVD and control cohorts, and cognitive effect sizes (see table S5). Results indicated that for every 1 year of difference in education between SVD and control groups, the effect size decreased (indicating superior performance of the control groups) by an estimated 0.24 standard deviations in the domain of executive function (95% CI: -0.38, -0.10; p=0.003), 0.28 standard deviations in the domain of visuospatial ability (95% CI: -0.46, -0.10; p=0.009), and 0.31 standard deviations in the domain of language (95% CI: -0.46, -0.16; p=0.001). Including difference in years of education as a predictor in these models increased I^2^ values by approximately 2% in the domains of processing speed and delayed memory, but reduced I^2^ values by approximately 13% in the domain of visuospatial ability and language, although I^2^ values remained large. Meta-regression models for attention and reasoning produced df<4, so their results were considered unreliable.

The majority of the meta-regression models assessing the influence of age on cognitive effect size produced df<4, therefore, we further investigated this by re-running meta-analysis models excluding studies in which SVD and control groups were not matched for age. In these analyses effect size estimates were similar to the initial meta-analysis models and all models remained significant. I^2^ values reduced in all domains except reasoning and visuospatial ability, but remained large (see table S6).

Study quality was rated on a scale from 0-8. The mean study quality score was 4.97 (median 5, range 2-8). As a sensitivity analysis, we re-ran meta-analysis models excluding all studies with a quality score <5. The magnitudes of pooled estimated effect sizes were comparable to those using the full meta-analysis dataset, and all models remained significant (see table S7). I^2^ values reduced by a small amount in the domains of executive function, visuospatial ability, attention and language, but increased in all other domains.

## Discussion

Based on 3229 individuals with SVD and 3679 control participants from 69 studies, our meta-analyses demonstrated that on average individuals with SVD perform more poorly than controls on cognitive tests in the domains of processing speed, executive function, delayed memory, visuospatial ability, reasoning, attention, and language. These findings support the notion that SVD-related cognitive impairment is global, affecting all examined cognitive domains, and mirrors the global effects of SVD seen on brain imaging^84,85^. This global cognitive impairment was present for cohorts with cerebrovascular, cognitive, and non-clinical presentations of SVD, although cohorts with a cognitive presentation of SVD had greater deficits in executive function, delayed memory, and visuospatial ability.

Our findings concur with those of a recent meta-analysis of 27 studies by Vasquez and Zakzanis^86^, which compared cognitive abilities of participants with vascular cognitive impairment without dementia (n=794) and control subjects (n=1750), finding deficits (from largest to smallest) in processing speed, immediate memory, delayed memory, general cognitive ability, language, executive function, visuospatial ability and working memory. Together with our meta-analysis, these results suggest that cognitive changes associated with SVD extend beyond impaired executive function and slowed processing speed to include multiple other domains, which often remain untested due to the perception that they are less affected in vascular cognitive impairment.

Results of our meta-regression analyses suggested that differences in years of education between SVD and control groups account for a proportion of the differences in cognitive test scores in the domains of memory, executive function, and visuospatial ability. All other cognitive domains showed a similar direction of effect (albeit non-significant) except processing speed, which could support the suggestion that processing speed might be less amenable to beneficial effects of education than other cognitive abilities^87^. A recent meta-analysis of early life risk factors in cerebrovascular disease found that fewer years of education was associated with increased MRI markers of SVD^88^. Similarly, our findings also highlight education as a (potentially modifiable) risk factor for SVD-related cognitive impairment, emphasising the importance of accounting for an individuals’ level of education in analyses of cognitive change over time, or comparisons of cognitive ability between groups.

An estimation of cognitive ability prior to the onset of decline is another potential confound in assessments of cognitive decline, as any change in cognitive ability will be relative to an individuals’ prior level^89^. Despite this, prior cognitive ability is seldom considered in clinical studies. Of the 69 studies included in our meta-analysis, only seven^15,19,21,49,50,60,68^ estimated prior cognitive ability using a test such as the National Adult Reading Test (NART), and only two of these studies included this score as a covariate in their analyses. As the NART is a vocabulary task, it was included as a cognitive outcome it in our meta-analysis of language. Therefore, our finding that individuals with SVD score more poorly on tests of language than control cohorts could reflect lower premorbid cognitive ability in the SVD cohorts, in addition to any decline in language abilities as a result of SVD.

A key strength of this study is that we did not pre-select literature that focuses on a certain lesion type, or clinical, cognitive, or behavioural presentation of SVD. Additionally, we included studies published in any language, which enabled us to analyse data from 18 countries in six continents. This broad approach to the characterisation of SVD aimed to capture a range of cohorts that represent the diversity of SVD presentations in different cultural and ethnic groups, and enable us to apply our findings to a range of clinical contexts. However, our study also has several important limitations. We observed high levels of heterogeneity in our meta-analyses. Whereas including SVD presentation and differences in demographic and vascular risk factors between SVD and control cohorts as predictors in meta-regression analyses reduced the I^2^ values of some models, we were unable to account for the vast majority of the heterogeneity we observed. One reason for this could be our use of group-level demographic and vascular risk data, which may limit the power to detect interactions between individual-level covariates and cognitive effect sizes. Meta-analytic approaches utilising individual patient data are increasingly popular, but rely upon the availability of patient-level datasets, which in our sample were rare. Incomplete reporting of vascular risk data also limited our assessment of their impact on cognitive effect sizes. Approximately half of all included studies reported history of hypertension and diabetes, but only one third of studies reported smoking status, despite its known association with SVD progression^1^. Similarly, we were unable to assess whether age at presentation to clinical services accounted for apparent differences in performance on memory tests between cohorts with a cognitive presentation of SVD and other SVD presentations - very few studies reported data on disease duration, with the exception of eight studies of stroke populations that reported average/minimum/maximum duration since stroke onset^21,33,61,64,65,70,75^. We were also limited by our reliance on the quality of study reporting; our literature search identified three studies whose results were inaccurately reported, or were statistically implausible and so, were excluded from our analyses^90–92^.

SVD-related cognitive impairment extends beyond deficits in processing speed and executive function - it affects a broader range of cognitive domains than previously considered. Our findings support the use of cognitive test batteries that cover a range of cognitive domains to fully investigate the extent of SVD-related cognitive impairment. Future investigations should include individuals with varying presentations of SVD, to represent the diversity of its clinical and non-clinical manifestations, and to enable more accurate characterisation and comparison of SVD subtypes. Accounting for educational level, or estimates of premorbid cognitive ability is essential for accurate assessment of SVD-related cognitive ability, and more complete reporting of vascular risk data will enable further exploration of the relative contributions of vascular risk factors to vascular cognitive impairment.

## Data Availability

The datasets analysed during the current study are available from the corresponding author on reasonable request.

## Contributors

The study was designed by OH and supervised by JMW and IJD. OH conducted the literature search and OH, EB, TR, AS, and CM screened papers for inclusion in the review. OH, EB, XL and EJ extracted data from eligible papers. OH and AJ categorised cognitive test data into domains. OH analysed and interpreted the data with contributions from JMW and IJD. OH wrote the first draft of the manuscript – all authors contributed to later versions.

## Acknowledgements

We would like to thank Rana Fetit, Noboru Komiyama, Masoud Shirali and Maria Valdéz Hernández, whose assistance with screening non-English texts for inclusion in the review was greatly appreciated. Thanks also Ezgi Tanriver Ayder and Francesca Chappell for their advice on statistical aspects of the meta-analysis. OH is funded by the College of Medicine and Veterinary Medicine at the University of Edinburgh, and is supported by the Wellcome Trust through the Translational Neuroscience PhD programme at the University of Edinburgh. JMW is supported by the Row Fogo Charitable Trust, Fondation Leducq (Perivascular Spaces Transatlantic Network of Excellence), and EU Horizon 2020 (SVDs@Target) and the MRC UK Dementia Research Institute at the University of Edinburgh. IJD is partly supported by grants that contribute to the Lothian Birth Cohorts (Age UK [Disconnected Mind grant], Medical Research Council [MR/R024065/1], and national Institutes of Health [1R01AG054628-01A1.)]). AS and TR are supported by funding from the Wellcome Trust 4-year PhD in Translational Neuroscience [108890/Z/15/Z].

